# A comparative multi-center study on the clinical and imaging features of confirmed and unconfirmed patients with COVID-19

**DOI:** 10.1101/2020.03.22.20040782

**Authors:** Congliang Miao, Jinqiang Zhuang, Mengdi Jin, Huanwen Xiong, Peng Huang, Qi Zhao, Li Miao, Jiang Du, Xinying Yang, Peijie Huang, Jiang Hong

**Author notes:** **Corresponding authors:** Jiang Hong, MD, PhD., Department of Internal and Emergency Medicine, Shanghai General Hospital, Shanghai Jiao Tong University, Xinsongjiang Road 650, Shanghai 201600, China. and Peijie Huang, MD.,Department of Internal and Emergency Medicine, Shanghai General Hospital, Shanghai.,Jiao Tong University.,Xinsongjiang Road 650, Shanghai 201600, China., Jiang Hong and Peijie Huang are Co-corresponding authors. Congliang Miao and Jinqiang Zhuang contributed equally to this work. **Conflict of Interest:** None declared.

## Abstract

**Background:** Previous studies had described the differences in clinical characteristics between ICU and non-ICU patients. However, seldom study focused on confirmed and unconfirmed groups. Our aim was to compare clinical and imaging characteristics of COVID-19 patients outside Hubei province between confirmed and unconfirmed group.

**Methods:** We retrospectively enrolled 163 consecutive adult patients with suspected COVID-19 from three tertiary hospitals in two provinces outside Hubei province from January 12, 2020 to February 13, 2020 and the differences in epidemiological, clinical, laboratory and imaging characteristics between the two groups were compared.

**Results:** This study enrolled 163 patients with 62 confirmed cases and 101 unconfirmed cases. Most confirmed patients were clustered (31, 50.0%) and with definite epidemiological exposure. Symptoms of COVID-19 were nonspecific, largely fever and dry cough. Laboratory findings in confirmed group were characterized by normal or reduced white blood cell count, reduced the absolute value of lymphocytes, and elevated levels of C-reactive protein (CRP) and accelerated Erythrocyte sedimentation rate (ESR). The typical chest CT imaging features of patients with confirmed COVID-19 were peripherally distributed multifocal GGO with predominance in the lower lung lobe. Compared with unconfirmed patients, confirmed patients had significantly higher proportion of dry cough, leucopenia, lymphopenia and accelerated ESR (*P*<0.05); but not with alanine aminotransferase, aspartate aminotransferase, D-dimer, lactic dehydrogenase, and myoglobin (*P*>0.05). Proportion of peripheral, bilateral or lower lung distribution and multi-lobe involvement, GGO, crazy-paving pattern, air bronchogram and pleural thickening in the confirmed group were also higher (*P*<0.05).

**Conclusions:** Symptoms of COVID-19 were nonspecific. Leukopenia, lymphopenia and ESR, as well as chest CT could be used as a clue for clinical diagnosis of COVID-19.

## Introduction

Since December 2019, a cluster of pneumonia cases with unknown etiology were reported in Wuhan, in the Hubei province of China(1). Scientists quickly identified the pathogen from infected patients respiratory epithelial cells and named it as severe acute respiratory syndrome corona virus 2 (SARS-Cov-2)(2). SARS-Cov-2 belongs to beta coronavirus and has 80% similar genetic sequence to severe acute respiratory syndrome (SARS)(3). SARS-Cov-2, as a pathogenic microorganism for a new infectious disease, is currently not well understood by scientists.

With the spread of the epidemic, 2019 coronavirus disease (COVID-19), the new disease caused by SARS-Cov-2, broke out nationwide and worldwide, especially China, Italy, South Korea, Iran. As to March 10, 2020, a total of 80924 laboratory-confirmed cases had been reported in China, and 32778 confirmed cases overseas, which represents a trend of global spread(4). On 11 March 2020, the world health organization declared the COVID-19 as a global pandemic. Theoretically, the most effective strategy for epidemic is early diagnosis, quarantine and treatment. Therefore, how to diagnose COVID-19 early is of great importance. It’s now the period of seasonal influenza, and the clinical symptoms of COVID-19 are similar to common respiratory diseases (influenza, common cold, viral pneumonia, etc.), which indicates the differential diagnosis are difficult for clinicians. At present, the diagnostic gold standard for COVID-19 is mainly viral nucleic acid reverse-transcription polymerase-chain-reaction (RT-PCR) testing, but considering its limited production, time consuming and relatively high false negative rate(5, 6), it might not effectively control the spread. Previous studies had described the differences in clinical characteristics between ICU and non-ICU patients(7-10), however very few scientists focused on confirmed and unconfirmed groups(11). Therefore, we described and compared the epidemiological history, clinical, laboratory characteristics and imaging features between confirmed and unconfirmed COVID-19 groups aim to providing a reference for the COVID-19 differential diagnosis, prevention and treatment.

## Methods

### Study Design and Participants

In this retrospective multi-center study, 163 consecutive and suspected adult patients were enrolled from three tertiary hospitals in two provinces outside Hubei province of China. Those patients who visited fever emergency clinics at Shanghai General Hospital, High-tech hospital (First Affiliated Hospital of Nanchang University) and People’s Hospital of Yinchun City from January 12, 2020 to February 13, 2020 were arranged for laboratory examinations and chest computed tomography (CT). Each suspected case was quarantined in separate rooms after consultation. Patients were transferred to a specialized hospital after diagnosis. This study was approved by the Ethics of Committees of Shanghai General Hospital Affiliated to School of Medicine of Shanghai Jiao Tong University. Informed consent for this retrospective study was waived.

### Patient and public involvement

No patient and public were involved in this study

### Data Collection

We retrospectively collected demographic data, medical history, epidemiological, clinical, laboratory, and CT imaging characteristics of all suspected patients on admission from medical records. Laboratory examinations consisted of blood routine, liver and renal function, electrolyte, coagulation testing, lactate dehydrogenase (LDH), creatine kinase, myoglobin, and troponin I, C-reactive protein (CRP), procalcitonin, erythrocyte sedimentation rate (ESR). We evaluated the presented imaging features (including lesion morphology and distribution features) of all patients with chest CT on admission.

### RT-PCR

Nasopharyngeal swabs or sputum specimens were collected and sent to the local center for disease control and prevention (CDC) for detection of SARS-Cov-2 by RT-PCR tests. Patients with first negative results should receive repeated tests in an interval of at least 1 day. We used forward primer, reverse primer and probe target to envelope gene of SARS-Cov-2. The sequence of forward primer was 5’-TCAGAATGCCAATCTCCCCAAC-3,’; The sequence of reverse primer was 5’-AAAGGTCCACCCGATACATTGA-3’; and The sequence of the probe was 5’CY5-CTAGTTACACTAGCCATCCTTACTGC-3’BHQ1. Conditions for the amplifications of the real-time RT-PCR were 50°C for 15 min, 95°C for 3 min, followed by 45 cycles of 95°C for 15 s and 60°C for 30 s.

### CT Review

All patients underwent chest CT scan in three hospitals of two provinces in China, 64 patients from Shanghai General Hospital were scanned on SOMATOM Definition Flash, Japan. 48 patients from High-tech hospital, First Hospital Affiliated of Nanchang University (Jiangxi Province) were scanned on Revolution Frontier, GE; And 51 patients from People’s Hospital of Yichun city (Jiangxi Province) were scanned on Optima 670 CT scanner, GE. Examination followed the normal chest protocols. CT scans were performed during end-inspiration with the patient in the supine position. Overall scan time is 2s, and slice thickness for reconstruction is 1.25 mm. All the reports were issued after two radiologists reviewed the CT images independently and would be concluded by a chief radiologist when opinions diverge.

### Study Definitions

All patients with suspected COVID-19 enrolled in this study were diagnosed according to WHO interim guidance(12). Nasopharyngeal swabs or sputum specimens were collected for RT-PCR tests. All the suspected cases were divided into confirmed and unconfirmed groups according to the results of real-time RT-PCR tests. The confirmed group was defined as a positive result of at least one RT-PCR tests for SARS-Cov-2. The unconfirmed group was defined as all results of RT-PCR tests were negative.

### Statistical Analysis

The data of patients were recorded by Epidata. Normally distributed continuous variables were described as mean ± SD (*x* ± *s*) and skewed distribution was expressed as the median (interquartile range). Continuous variables were compared using independent t-test if they are normally distributed; otherwise, the Mann-Whitney test was used. Categorical variables were described as frequency rates and percentages. Proportions for categorical variables were compared using the chi-square test or the Fisher exact test. All statistical analyses were performed using SPSS version 13.0 software (SPSS Inc). All statistical tests were 2-sided, and *P*<0.05 was regarded as statistically significant.

## Results

### Demographic, epidemiological and clinical characteristics

This study enrolled 163 patients with 62 cases confirmed and 102 cases unconfirmed according to RT-PCR test results. The demographic, epidemiological and clinical characteristics of patients were shown in the Table 1. The mean age of patients in confirmed group was 43.8±13.9 years (19-77), and 32 (51.6%) were men. Of these patients, 4 cases (6.5%) were critically illness and 3 (4.8%) patients with asymptomatic infection. 8 (12.9%) of the patients in confirmed group had coexisting chronic diseases. The average time from epidemiological exposure to symptoms onset was 7.0 (3.5-9.0) days. A total of 31 (50.0%) patients were clustered and 2 patients had a history of exposure to the Huanan seafood market; 40 patients (71%) were residents of Wuhan or had visited the city; 29 (46.8%) cases with contact confirmed COVID-19 or febrile patients; exposure history was unknown in 12 (19.4%) patients. The most common symptoms on admission were fever (49 [79.0%]), dry cough (37 [59.7%]), fatigue or myalgia (15 [24.2%]). Less common symptoms were productive cough, diarrhea, chest tightness, headache, sore throat, upper airway congestion, dyspnea. In the unconfirmed patients, the average age was 41.3±14.7 years (range 19-81 years), with 68 males (67.3%). Compared with patients in unconfirmed group, patients in confirmed group had significantly higher proportion of Wuhan residence history, having visited Wuhan, clustering diseases and dry cough. It was found that proportion of males and productive cough in the confirmed group were less than those in the unconfirmed group (*P*<0.05), while the remaining variables were not significant (*P*>0.05).

**Table 1.** General demographic, epidemiological and clinical characteristics between confirmed group and unconfirmed group

| Characteristics | Unconfirmed group<br>n=101 | Confirmed group<br>n=62 | P value |
| --- | --- | --- | --- |
| Age, years | 41.3±14.7 | 43.8±13.9 | 0.28 |
| Sex |  |  |  |
| Men, n(%) | 68(67.3%) | 32(51.6%) | <0.05 |
| Women, n(%) | 33(32.7%) | 30(48.4%) |  |
| Systolic pressure, mmHg | 120.8±10.9 | 121.5±12.2 | 0.76 |
| Diastolic pressure, mmHg | 74.9±8.8 | 75.4±9.3 | 0.76 |
| Heart rate, bpm | 83.0(75.8-88.0) | 79.5(74.8-85.0) | 0.10 |
| Respiratory rate, bpm | 19(16-20) | 19.5(18.0-20.0) | 0.64 |
| SaO<sub>2</sub>, % | 97(95-98) | 98(96-99) | 0.20 |
| Symptoms onset to the first visit, day(s) | 2.0(1.0-5.0) | 3.0(1.0-6.0) | 0.39 |
| Exposure to symptoms onset, day(s) | 6.0(4.0-9.0) | 7.0(3.5-9.0) | 0.92 |
| Smoking, n(%) | 25(24.8%) | 11(17.7%) | 0.30 |
| Drinking, n(%) | 17(16.8%) | 8(12.9%) | 0.50 |
| **Epidemiological history** |  |  |  |
| Patients with Wuhan residence history, n(%) | 14(13.9%) | 22(35.5%) | <0.01 |
| Patients with Wuhan travel history, n(%) | 30(29.7%) | 22(35.5%) | <0.01 |
| Huanan seafood market exposure, n(%) | 0(0%) | 2(3.2%) | 0.14 |
| Contact with patients of fever or COVID-19, n(%) | 39(38.6%) | 29(46.8%) | 0.31 |
| Clustering occurrence, n(%) | 11(10.9%) | 31(50.0%) | <0.01 |
| **Any comorbidity** |  |  |  |
| History of hypertension, n(%) | 14(13.9%) | 7(11.3%) | 0.63 |
| History of diabetes, n(%) | 5(5.0%) | 3(4.8%) | 1.00 |
| History of cardiovascular disease, n(%) | 2(2.0%) | 1(1.6%) | 1.00 |
| History of cerebrovascular disease, n(%) | 1(1.0%) | 1(1.6%) | 1.00 |
| Chronic obstructive pulmonary disease, n(%) | 2(2.0%) | 0(0%) | 0.53 |
| History of chronic liver disease, n(%) | 3(3.0%) | 0(0%) | 0.29 |
| History of chronic kidney disease, n(%) | 0(0%) | 0(0%) | - |
| History of cancer, n(%) | 1(1.0%) | 0(0%) | 1.00 |
| **Clinical symptoms** |  |  |  |
| Fever, n(%) | 77(76.2%) | 49(79.0%) | 0.68 |
| Highest temperature, °C | 37.7±0.8 | 37.7±0.7 | 0.56 |
| Dry cough, n(%) | 38(37.6%) | 37(59.7%) | <0.01 |
| Sputum production, n(%) | 39(38.6%) | 10(16.1%) | <0.01 |
| Myalgia or fatigue, n(%) | 16(15.8%) | 15(24.2%) | 0.19 |
| Sore throat, n(%) | 10(9.9%) | 5(8.1%) | 0.69 |
| Headache, n(%) | 9(8.9%) | 5(8.1%) | 0.85 |
| Diarrhea, n(%) | 4(4.0%) | 7(11.3%) | 0.11 |
| Upper airway congestion, n(%) | 9(8.9%) | 3(4.8%) | 0.33 |
| Chest tightness, n(%) | 14(13.9%) | 6(9.7%) | 0.43 |
| Dyspnea, n(%) | 5(5.0%) | 3(4.8%) | 1.00 |
| Critically ill patients, n(%) | 2(2.0%) | 4(6.5%) | 0.20 |
| Asymptomatic infection, n(%) | 4(4.0%) | 3(4.8%) | 1.00 |

COVID-19, 2019 coronavirus disease

### Laboratory Parameters between Confirmed group and Unconfirmed group

The laboratory findings of patients are summarized in the Table 2. On admission, lymphocytopenia was present in 35.5% of the confirmed group, and leukopenia in 24.2%. Most of the confirmed patients had elevated levels of CRP and ESR. Compared with unconfirmed group, confirmed group had significantly lower white blood cell count, neutrophil count and the absolute value of lymphocytes. It also had higher proportion of leukopenia and lymphopenia. ESR and potassium on admission were higher in confirmed group than unconfirmed group, while other laboratory findings did not significantly differ between groups (*P*>0.05).

**Table 2.** Laboratory Parameters between confirmed group and unconfirmed group

| Variables | Unconfirmed group<br>n=101 | Confirmed group<br>n=62 | P value |
| --- | --- | --- | --- |
| White blood cell count, $\times 10^9/L$ | 7.6(6.0-9.1) | 5.3(4.0-6.7) | <0.01 |
| White blood cell count($\times 10^9/L$), n(%) | | | <0.01 |
| <4 | 7(6.9%) | 15(24.2%) |  |
| 4-10 | 79(78.2%) | 43(69.4%) |  |
| $\geq 10$ | 15(14.9%) | 4(6.5%) | |
| Neutrophil count, $\times 10^9/L$ | 5.1(3.7-6.6) | 2.9(2.3-4.3) | <0.01 |
| Lymphocyte count, $\times 10^9/L$ | 1.7(1.2-2.2) | 1.3(0.9-1.8) | <0.01 |
| Lymphocyte count<$1 \times 10^9/L$, n(%) | 19(18.8%) | 22(35.5%) | 0.02 |
| Hemoglobin, g/L | 144.9±19.9 | 138.8±16.7 | 0.05 |
| Platelet count, $\times 10^9/L$ | 199.0(166.5-247.0) | 178.0(130.5-215.5) | 0.01 |
| Erythrocyte sedimentation rate | 15.0(10.5-28.5) | 26.0(15.3-49.8) | 0.04 |
| C-reactive protein, mg/L | 10.9(1.5-30.3) | 9.9(3.3-26.2) | 0.74 |
| Procalcitonin, ng/ml | 0.05(0.03-0.06) | 0.05(0.05-0.05) | 0.78 |
| Procalcitonin(ng/ml), n(%) |  |  | 1.00 |
| <0.1 | 98(97.0%) | 61(98.4%) |  |
| $\geq 0.1$ | 3(3.0%) | 1(1.6%) | |
| Alanine aminotransferase, U/L | 25.0(16.7-37.0) | 22.0(15.5-34.5) | 0.36 |
| Alanine aminotransferase(U/L), n(%) |  |  | 0.91 |
| $\leq 40$ | 84(83.2%) | 52(83.9%) | |
| >40 | 17(16.8%) | 10(16.1%) |  |
| Aspartate aminotransferase, U/L | 24.0(21.0-33.8) | 25.5(20.0-30.8) | 0.98 |
| Total bilirubin, mmol/L | 8.9(7.0-12.8) | 9.7(7.4-13.1) | 0.56 |
| Lactate dehydrogenase, U/L | 239.0(177.0-403.2) | 208.0(170.3-265.0) | 0.13 |
| Lactate dehydrogenase(U/L), n(%) |  |  | 0.14 |
| $\leq 245$ | 62(61.4%) | 45(72.6%) | |
| >245 | 39(38.6%) | 17(27.4%) |  |
| Creatine kinase, U/L | 91.0(63.0-122.0) | 67.0(53.4-105.8) | 0.04 |
| Creatine kinase(U/L), n(%) |  |  | 1.00 |
| $\leq 185$ | 95(94.1%) | 58(93.5%) | |
| >185 | 6(5.9%) | 4(6.5%) |  |
| Myoglobin, ng/ml | 22.2(13.7-52.0) | 9.0(2.9-19.1) | 0.03 |
| Troponin I, ng/ml | 0.01(0.00-0.01) | 0.01(0.00-0.01) | 0.61 |
| Troponin I(ng/ml), n(%) |  |  | 1.00 |
| $\leq 0.04$ | 98(97.0%) | 60(96.8%) | |
| >0.04 | 3(3.0%) | 2(3.2%) |  |
| Potassium, mmol/L | 3.9(3.6-4.0) | 4.1(3.8-4.3) | <0.01 |
| Sodium, mmol/L | 138.6(137.0-140.0) | 138.4(136.0-139.4) | 0.11 |
| Creatine, $\mu\text{mol/L}$ | 70.0(55.0-84.3) | 68.0(56.0-76.6) | 0.33 |
| D-dimer, mg/L | 0.29(0.19-0.54) | 0.37(0.26-0.74) | 0.12 |
| Prothrombin time, Sec | 11.4(11.1-12.9) | 11.4(11.0-11.9) | 0.36 |
| Activated partial thromboplastin time, Sec | 28.4±3.9 | 29.1±5.7 | 0.50 |

### Comparisons in CT imaging features between Confirmed group and Unconfirmed group

The CT imaging features on admission are shown in Table 3. On admission, 9 patients did not have CT imaging abnormality in confirmed group. In confirmed group, 42 (67.7%) patients had bilateral lung involvement, 34 (54.8%) showed peripheral distribution, and 46 (74.2%) showed more than 2 affected lobes. Two lobes and all lobes involved were more common, with 18 cases (29.0%) and 13 cases (21.0%) respectively. Compared with unconfirmed patients, confirmed patients had significantly higher proportion of peripheral, bilateral or lower lung distribution and multi-lobe involvement (*P*<0.05). The most common patterns seen on chest CT in confirmed group were ground-glass opacity (GGO) (39 cases, 62.9%), followed by crazy-paving pattern (17 cases, 27.4%) and air bronchogram (14 cases, 22.6%), then consolidation (12 cases, 19.4%) and pleural thickening (12 cases, 19.4%) (Figure 1). Cavitation, pleural effusion, lymphadenopathy and pulmonary fibrosis were rare. Proportion of GGO, crazy-paving pattern, air bronchogram, and pleural thickening in the confirmed group were also higher (*P*<0.05). The remaining variables were not (*P*>0.05).

**Table 3.**
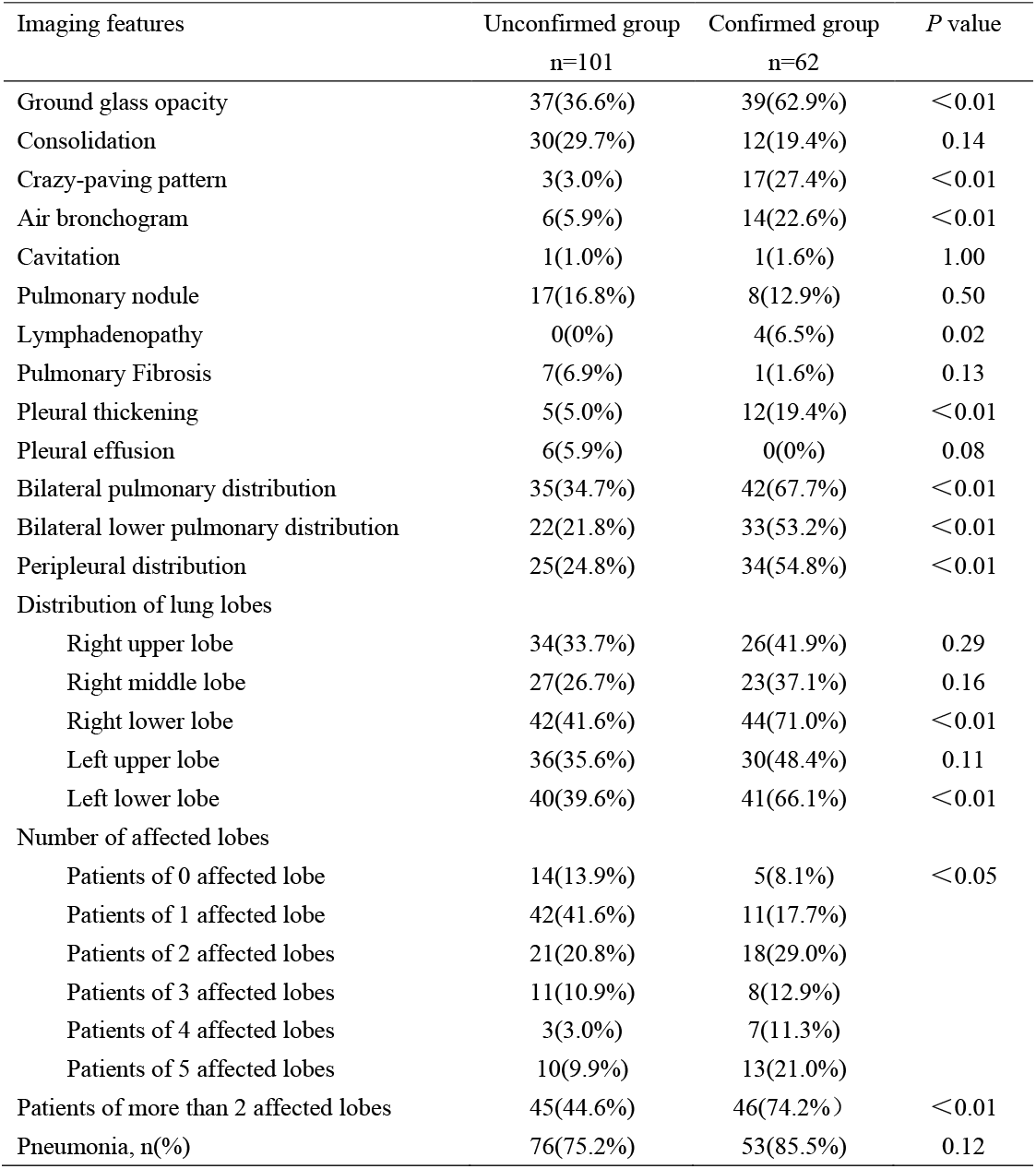
Comparisons of CT imaging features between confirmed group and unconfirmed group

| Imaging features | Unconfirmed group<br>n=101 | Confirmed group<br>n=62 | *P* value |
| --- | --- | --- | --- |
| Ground glass opacity | 37(36.6%) | 39(62.9%) | <0.01 |
| Consolidation | 30(29.7%) | 12(19.4%) | 0.14 |
| Crazy-paving pattern | 3(3.0%) | 17(27.4%) | <0.01 |
| Air bronchogram | 6(5.9%) | 14(22.6%) | <0.01 |
| Cavitation | 1(1.0%) | 1(1.6%) | 1.00 |
| Pulmonary nodule | 17(16.8%) | 8(12.9%) | 0.50 |
| Lymphadenopathy | 0(0%) | 4(6.5%) | 0.02 |
| Pulmonary Fibrosis | 7(6.9%) | 1(1.6%) | 0.13 |
| Pleural thickening | 5(5.0%) | 12(19.4%) | <0.01 |
| Pleural effusion | 6(5.9%) | 0(0%) | 0.08 |
| Bilateral pulmonary distribution | 35(34.7%) | 42(67.7%) | <0.01 |
| Bilateral lower pulmonary distribution | 22(21.8%) | 33(53.2%) | <0.01 |
| Peripleural distribution | 25(24.8%) | 34(54.8%) | <0.01 |
| Distribution of lung lobes |  |  |  |
| Right upper lobe | 34(33.7%) | 26(41.9%) | 0.29 |
| Right middle lobe | 27(26.7%) | 23(37.1%) | 0.16 |
| Right lower lobe | 42(41.6%) | 44(71.0%) | <0.01 |
| Left upper lobe | 36(35.6%) | 30(48.4%) | 0.11 |
| Left lower lobe | 40(39.6%) | 41(66.1%) | <0.01 |
| Number of affected lobes |  |  |  |
| Patients of 0 affected lobe | 14(13.9%) | 5(8.1%) | <0.05 |
| Patients of 1 affected lobe | 42(41.6%) | 11(17.7%) |  |
| Patients of 2 affected lobes | 21(20.8%) | 18(29.0%) |  |
| Patients of 3 affected lobes | 11(10.9%) | 8(12.9%) |  |
| Patients of 4 affected lobes | 3(3.0%) | 7(11.3%) |  |
| Patients of 5 affected lobes | 10(9.9%) | 13(21.0%) |  |
| Patients of more than 2 affected lobes | 45(44.6%) | 46(74.2%) | <0.01 |
| Pneumonia, n(%) | 76(75.2%) | 53(85.5%) | 0.12 |

**Figure 1.**
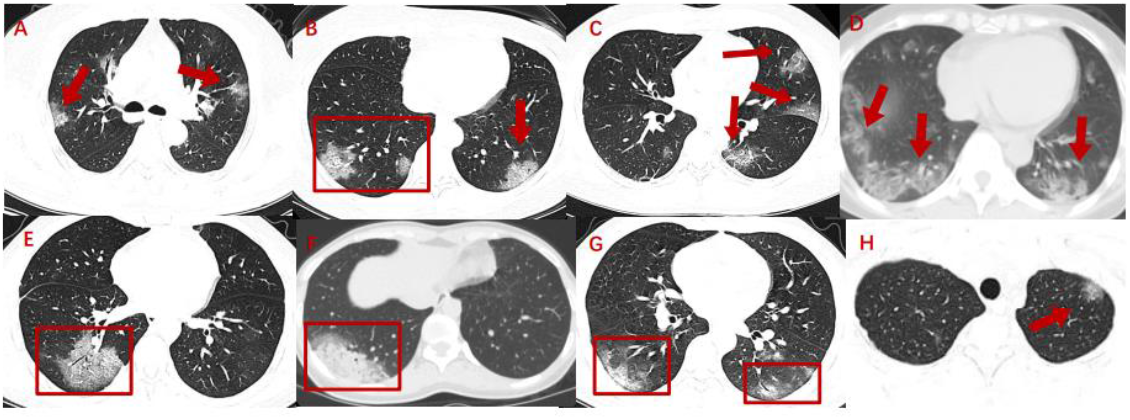
Various chest CT imaging features of COVID-19 GGO, ground glass opacity A, GGO with bilateral and subpleural distribution; B, crazy-paving pattern with subpleural and bilateral lower pulmonary distribution; C, GGO; D, GGO with bilateral and subpleural distribution; E, GGO with crazy-paving pattern and air bronchogram; F, GGO with consolidation; G, GGO with air bronchogram and subpleural distribution; H, GGO with unilateral and subpleural distribution.

## Discussion

This study showed that most patients with COVID-19 had pneumonia, and the most common symptoms at onset of illness were fever, dry cough, myalgia or fatigue. Less common symptoms were diarrhea, headache, sore throat, upper airway congestion and a small number of severe patients developed chest tightness and dyspnea, indicating that the target cells might be located in the lower airway. This was consistent with the results of previous studies(7-10, 13-15). There were no significant differences in initial clinical symptoms between in Hubei province and outside Hubei province. These nonspecific symptoms are shared by winter respiratory diseases caused by bacteria and viruses, especially influenza. Influenza are usually characterized by fever, myalgia, headache and dry cough, most of which are self-limiting but may also progress to severe conditions, such as pneumonia, myocarditis and death(16, 17). In addition, severe acute respiratory syndrome corona virus (SARS-CoV) and Middle East respiratory syndrome corona virus (MERS-CoV) also caused similar clinical symptoms, but more frequent diarrhea symptoms(18, 19). To the best of our knowledge, this was the first multi-center study to compare the clinical features between confirmed group and unconfirmed group. This report demonstrated that the incidence of dry cough in confirmed group was significantly higher than that in unconfirmed group, but the clinical symptoms of patients with COVID-19 were nonspecific. Therefore, clinical symptoms alone were difficult to distinguish from winter respiratory diseases.

Our study showed that the white blood cell count and the absolute value of lymphocytes in confirmed group was significantly lower than that in unconfirmed group. Qin et al demonstrated that lymphocytes counts in most severe COVID-19 patients was reduced, which suggests that SARS-CoV2 might impaired immune system and mainly damaged lymphocytes, especially T lymphocytes(20). This was similar to SARS-CoV, which spread through the respiratory mucosa and infect other cells, triggering a series of inflammatory responses and cytokine storm, resulting in changes in lymphocytes(21). Therefore, the reduced white blood cell count and lymphocytes could be used as a reference index for clinical diagnosis of COVID-19. In addition, the ESR of the confirmed group was significantly higher than that of unconfirmed group, while there was no statistical difference in CRP and procalcitonin between the two groups. Therefore, ESR could help clinicians distinguish between COVID-19 and winter respiratory diseases. Although previous studies had shown that some patients with confirmed COVID-19 had abnormal alanine aminotransferase (ALT), aspartate aminotransferase (AST), LDH, D-dimer, myoglobin and troponin I, but there was no statistical significance between the two groups in this study. The underlying reason might be that the majority of patients with COVID-19 outside Hubei province were mild infections, with a small proportion of critically ill cases, while hepatic injury,\ coagulation activation, myocardial injury were common in critically ill and severe cases. This result was contrary to a recent study. Zhao et al showed that the levels of liver function associated markers (ALT, AST and LDH) were significantly higher in confirmed group than in unconfirmed group, but the sample size of the study was very small(11). Therefore, these laboratory indicators might not provide reference for the differential diagnosis of COVID-19 outside Hubei province.

The CT imaging features of COVID-19 are diversity. SARS-Cov-2 belongs to β coronavirus family, which causes pneumonia with similar imaging features to other coronavirus pneumonia, such as SARS and Middle East respiratory syndrome (MERS)(22, 23). Our study showed that the typical CT imaging features of COVID-19 were peripherally distributed multifocal GGO with predominance in the lower lung lobe. However, it still had specific imaging characteristics, especially compared to that of influenza A and influenza B. While the manifestations of seasonal influenza are small patch GGO and consolidation with subpleural and/or peribronchial distribution on chest CT(24). Our study indicated that the proportion of GGO, crazy-paving pattern, air bronchogram, and pleural thickening in the confirmed group was significantly higher than that in the unconfirmed group. In terms of the distribution of lesions, the proportion of bilateral, subpleural, lower lung distribution and multi-lobe involvement in the confirmed group was significantly higher than that of the unconfirmed group. Considering the high incidence of seasonal influenza, a recent study showed that the most common etiology in all suspected cases was influenza virus infection(25). Therefore, chest CT could become an effective clinical diagnostic tool for screening patients with suspected COVID-19. Previous studies had demonstrated the high sensitivity of chest CT in the diagnosis of COVID-19 and emphasized the important role of chest CT in early diagnosis(6, 26). In this study, 4 cases with asymptomatic infections of the 62 confirmed COVID-19 patients were first detected by chest CT scan and finally confirmed by RT-PCR. Therefore, it could not only be possible to identify patients with negative results of RT-PCR but highly suspicious of COVID-19 for clinicians, but also to screen patients with a clear history of epidemiological exposure but asymptomatic infection. In addition, chest CT was helpful for monitoring the dynamic therapeutic effect of patients during treatment(27). It was worth noting that 9 of the 62 confirmed COVID-19 patients in this study had no abnormal lesions in chest CT. Therefore, suspected cases with epidemiological history, clinical manifestations and abnormal laboratory tests should be diagnosed combined with the results of RT-PCR test.

Our advantage was that this was the first multi-center study to compare the clinical and imaging features between confirmed and unconfirmed groups. Comparison of the clinical and imaging features between the two groups may help clinicians to differentiate COVID-19 from other pathogen infections and then to screen patients with highly suspected cases.

There are several limitations in our study. First, although the RT-PCR test is the current gold standard for the diagnosis of COVID-19, it might still present a certain false-negative rate after repetitions. This is mainly due to the fact that the test samples are mostly pharynx swabs rather than bronchoalveolar lavage fluid (BALF) and RNA was easily degraded. Second, we did not complete the tests of influenza A virus, influenza B virus, parainfluenza virus, mycoplasma pneumoniae, chlamydia pneumoniae, respiratory syncytial virus, adenovirus, coxsackie virus, the nucleic acid of influenza viruses A and B and microbial culture in all patients. We could not clearly distinguish the specific pathogen infections in patients with unconfirmed group. Therefore, we did not further compare the differences between COVID-19 and specific pathogen infections.

In summary, most patients with COVID-19 had a definite epidemiological history of exposure in Wuhan or to infected patients. The clinical symptoms of COVID-19 were nonspecific, largely fever and dry cough. The reduced white blood cell count, lymphocytes count and ESR could be used as a reference index for clinical diagnosis of COVID-19. Chest CT could become an effective clinical diagnostic tool for screening patients with suspected COVID-19, but the final diagnosis still needs to be combined with the results of RT-PCR tests.

## Data Availability

Raw data is only available if reasonably required

## Acknowledgements

None.

## Contributors

Congliang Miao and Jinqiang Zhuang contributed to the acquisition and analysis of the data and the initial writing of the draft of this paper. Peng Huang, Xinying Yang, Li Miao and Huanwen Xiong contributed to the collection and interpretation of data. Mengdi Jin and Jiang Du contributed to revise draft critically for important intellectual contents. Peijie Huang and Jiang Hong contributed to the concept and design of the study and the final approval of the version to be published.

## Funding information

The authors state that this work has not received any funding.

### Ethics approval

The study protocol was approved by the Ethics of Committees of Shanghai General Hospital Affiliated to School of Medicine of Shanghai Jiao Tong University.

### Patient consent for publication

Not required.

### Provenance and peer review

Not commissioned; externally peer reviewed.

### ORCID iDs

Jiang Hong http://orcid.org/0000-0002-3745-5549

